# Model-Based Assessment of Photoplethysmogram Signal Quality in Real-Life Environments

**DOI:** 10.1101/2024.06.07.24308621

**Authors:** Yan-Wei Su, Chia-Cheng Hao, Gi-Ren Liu, Yuan-Chung Sheu, Hau-Tieng Wu

## Abstract

Assessing signal quality is crucial for photoplethysmogram analysis, yet a precise mathematical model for defining signal quality is often lacking, posing challenges in the quantitative analysis. To tackle this problem, we propose a Signal Quality Index (SQI) based on the adaptive non-harmonic model (ANHM) and a Signal Quality Assessment (SQA) model, which is trained using the boosting learning algorithm. The effectiveness of the proposed SQA model is tested on publicly available databases with experts’ annotations. Result: The DaLiA database [20] is used to train the SQA model, which achieves favorable accuracy and macro-F1 scores in other public databases (accuracy 0.83, 0.76 and 0.87 and macro-F1 0.81, 0.75 and 0.87 for DaLiA-testing dataset, TROIKA dataset [31], and WESAD dataset [23], respectively). This preliminary result shows that the ANHM model and the model-based SQI have potential for establishing an interpretable SQA system.

## 1. Introduction

Photoplethysmogram (PPG) is widely utilized in clinical and consumer devices for their non-invasive and cost-effective nature [1]. Initially employed to measure blood oxygen saturation and monitor resting heart rate (HR), the PPG signal also holds rich information on the cardiovascular, respiratory, autonomic nervous systems, or even blood pressure, which has not been routinely exploited but has started to gain attention in the digital health era. However, like other biomedical signals, PPG information’s accuracy relies on signal quality, which is high at rest but usually diminishes with movement [2, 10]. Therefore, a robust signal quality assessment method is crucial to identify noise-corrupted segments, ensuring reliable measurements of parameters like heart rate and oxygen saturation from high-quality signal segments [19].

Various methods exist for assessing the quality of a PPG signal [15] under different criteria, such as the presence of clear pulse peaks [19, 8] for HR extraction or clean pulse waveforms, cardiac component, or visible systolic and diastolic waves [16] for diagnosis demands. Additional considerations include pulse amplitude and width consistency with adjacent pulses, adherence to typical PPG pulse morphology [28]. Alternatively, simultaneous recording of other signals, as demonstrated in [17], can be employed to define quality. To automatically quantify PPG quality, signal processing techniques are needed. This can be achieved through time-domain, frequency-domain, or hybrid approaches, guided by predefined rules or machine learning techniques. See [19] for a review. To our knowledge, experts seem to rely on the visibility of the cardiac component to label the quality of a PPG segment. Despite implicitly consented PPG signal quality criteria among experts and numerous proposed PPG signal quality assessments (SQA), there is, to our knowledge, no precise definition of PPG signal quality with a mathematical model, particularly in a free-living environment.

We model the PPG signal by the *adaptive non-harmonic model* (ANHM), which incorporates respiration-induced intensity variation (RIIV) [25] and motion rhythm, being non-sinusoidal when exists and apply time-frequency (TF) analysis to recover harmonics of the cardiac component. Based on these, we introduce our model-based signal quality index (SQI) to evaluate the quality of *cardiac component* residing in a PPG signal and an interpretable learning based SQA model based on the proposed and existing signal quality indices. We apply the SQA model to publicly available databases with expert annotations, showcasing its applicability.

## 2. Mathematical model and signal decomposition

It is well known that a PPG signal is composed of possible multiple components, including a cardiac component, a respiratory component [25], and a motion rhythm when a subject is exercising. The oscillatory morphology of the cardiac component changes from cycle to cycle, encoding the underlying physiological status [7], and the similar observation might hold for other components. Also, noise is inevitable. Jointly, we consider the *adaptive non-harmonic model* (ANHM) [13] to model the PPG signal. Fix small constants *ϵ,ϵ*^*′*^ > 0 and Δ > 0. We model a clean PPG signal by the ANHM [13]:

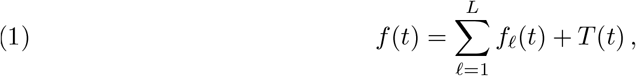

where

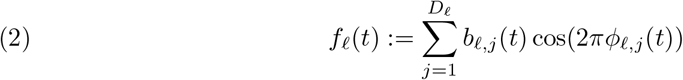

is called the *intrinsic model type* (IMT) function, *ϕ*_*𝓁*,1_ and 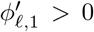 are called the *phase* and the *instantaneous frequency* (IF) of the *𝓁*-th IMT function, *b*_*𝓁,j*_(*t*) > 0 is the *amplitude modulation* (AM), and *T* (*·*) is a smooth function so that its Fourier transform 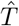 is compactly supported in [−Δ, Δ]. For each *𝓁* ∈ {1, …, *L*}, we assume the following additionally:

(C1) *ϕ*_*𝓁,j*_ ∈ *C*^2^(ℝ) for *j* = 1, …, *D*_*𝓁*_. When 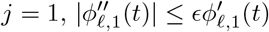 for all *t* ∈ ℝ; when 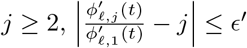 and 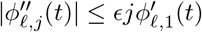 for all *t* ∈ ℝ.
(C2) *b*_*𝓁,j*_ ∈ *C*^1^(ℝ) for *j* = 1, …, *D*_*𝓁*_. When *j* ≥ 2, *b*_*𝓁,j*_(*t*) ≤ *c*_*𝓁,j*_*b*_*𝓁*,1_(*t*) and 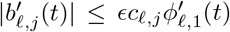 for all *t* ∈ ℝ, where *c*_*𝓁*,1_ > 0, *c*_*𝓁,j*_ ≥ 0 and 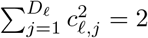
(C3) When *L* > 1, for any *t* ∈ ℝ, 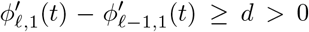 for *𝓁* = 2, …, *L*, and 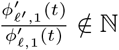 for any *𝓁 < 𝓁*^*𝓁*^.

When *ϵ*′ = 0 and *b*_*𝓁,j*_(*t*) = *c*_*𝓁,j*_*b*_*𝓁*,1_(*t*) for all *j* ≥ 2, the ANHM can be expressed as a function with *fixed* wave-shape functions (WSF); that is,

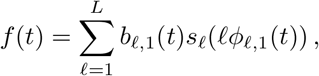

where *s*_*𝓁*_ is a 1-periodic function [13]. For each *𝓁* ∈ {1, …, *L*}, *D*_*𝓁*_ ∈ ℕ is called the *harmonic order* for the *𝓁*-th IMT function. When *D*_*𝓁*_ = 1, the *𝓁*-th IMT function oscillates with a sinusoidal WSF. Note that in general it is possible that

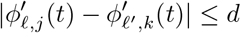

for *𝓁* ≠ *𝓁*′ and some *j, k* ∈ ℕ. We call *b*_*𝓁*,1_(*t*) cos(2*πϕ*_*𝓁*,1_(*t*)) the *fundamental component* of the *𝓁*-th IMT function, and for *j* > 1, we call *b*_*𝓁,j*_(*t*) cos(2*πϕ*_*𝓁,j*_(*t*)) the *j-th harmonic* of the *𝓁*-th IMT function. We refer readers to [13] for more detailed discussion of the model and these conditions. When *L* = 1, the only IMT function is the cardiac component, which *usually* can be well modeled by *D*_1_ = 6. When respiration and/or walking patterns exist, *L* > 1, and their harmonic orders are lower, like 3. In a PPG example shown in Figure 5, it is difficult to visualize the cardiac oscillation in the raw signal, even if it exists and is of high quality after decomposition. In practice, we can remove the trend component *T* by applying a high-pass filter, so from now on we assume *T* = 0.

With the ANHM model, we consider the *time-frequency* (TF) analysis approach to decompose the signal, due to the time-varying frequency and amplitude nature of PPG signals. This approach has been applied to solve several signal processing problems, such as the extraction of the phase and the amplitude information, signal decomposition into essential components (IMT functions and their harmonics), denoising, and dynamic feature extraction. While there are several choices, we consider the short-time Fourier transform (STFT) based synchrosqueezing transform (SST) [5, 18]. SST generates a TF representation (TFR) of the PPG signal. It has been theoretically established that when a signal adheres ANHM with sinusoidal WSFs, the ridges of STFT closely approximate the IFs of all IMT functions [6], and SST utilizes the phase information of STFT to sharpen the TFR and hence the performance of *ridge detection* (RD) is enhanced [14]. When we decompose a signal, we assume the knowledge of *L*. In general, estimating *L* is still a challenging problem, but estimating *D*_*𝓁*_ could be achieved by the trigonometric regression [22]. Under the ANHM, by the RD and reconstruction formula for SST [5], we could robustly and accurately estimate 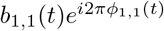 [3], and the first IMT function can be reconstructed via taking the real part of the superposition of these estimated harmonics components. By subtracting the first IMT function from the PPG signal, we proceed with reconstructing the second IMT function by the same approach. By iteration, we obtain a decomposition of all IMT functions. The overall flowchart of ridge detection and harmonic decomposition algorithm, shape-adaptive mode decomposition (SAMD) is shown in Figure 1.

**Figure 1.**
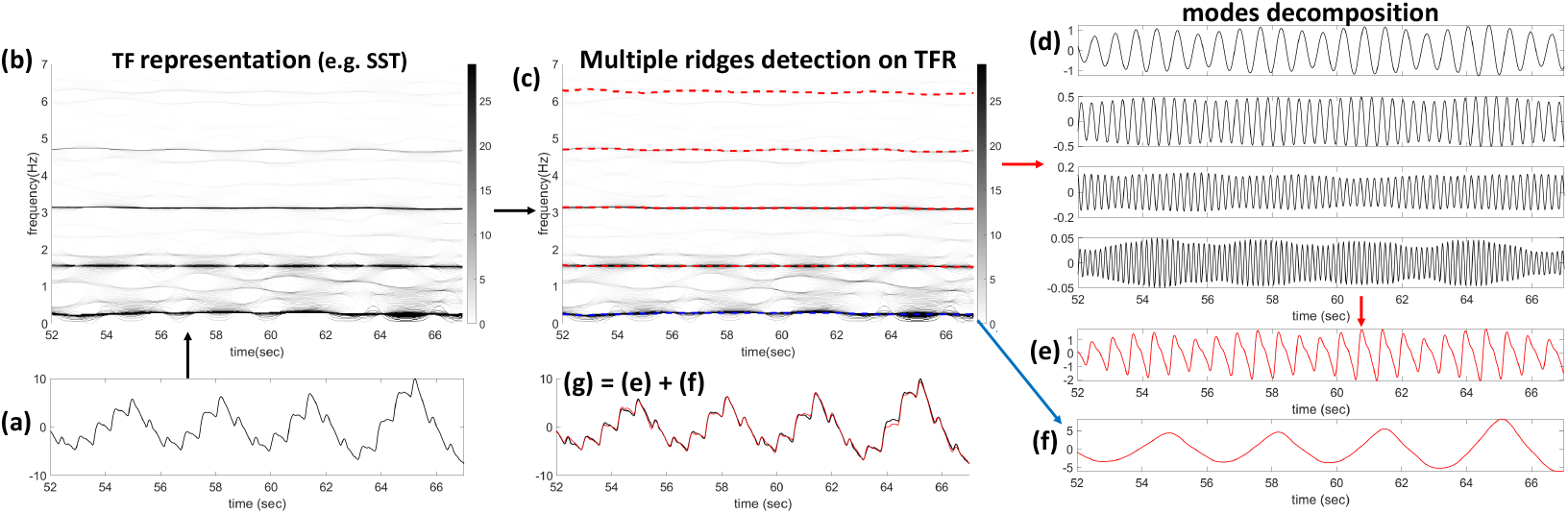
The overall flowchart of ridge detection and harmonic decomposition algorithm. (a) A segment of PPG signal that contains a cardiac component and a respiratory component. (b) The time-frequency representation of (a) determined by the second-order SST. (c) The detected ridges are superimposed as red-dashed curves on the TFR shown in (b). (d) The reconstructed harmonics of the cardiac component, which are related to the detected ridges shown in (c). (e) The reconstructed cardiac component, which comes from the superposition of reconstructed harmonics shown in (d). (f) The reconstruct fundamental component of the respiratory component that is related to the ridge shown as the blue-dashed curve in (c). (g) The superposition of (e) and (f).

## 3. Signal quality indices for a ppg signal

To our understanding, “signal quality” is a broad term typically described and quantified by *implicitly* equating it with the visibility of the cardiac component, sometimes considering conditions like systolic or diastolic phase behavior as indicators of high quality. Let us now quantify this traditional idea. To quantify this idea precisely, we model a PPG signal by (1), assume *𝓁* = 1 is the cardiac component, and define the SQI by

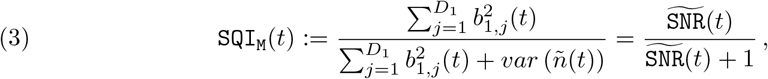

where

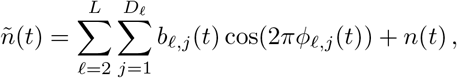

*n*(*t*) is the inevitable noise, and

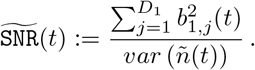

In other words, we view non-cardiac component as “noise”. Clearly, when the cardiac component as the signal is strong and the noise is weak, 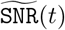 is large and `SQI`_M_ (*t*) is close to 1. Otherwise it is close to 0. In the *ideal* situation when *L* = 1, *ñ*(*t*) = *n*(*t*) and

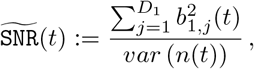

which is the relationship between the cardiac component and the noise.

In practice, the PPG signal is uniformly sampled at a fixed sampling rate, *f*_*s*_ Hz, and saved as a vector **x** ∈ ℝ^*N*^ ; that is,

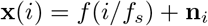

for 1 ≤ *i* ≤ *N*, where **n**_*i*_ is a mean zero noise with finite variance. Clearly, each component of *f* is also uniformly sampled as an ℝ^*N*^ vector. For example, the cardiac component is given by

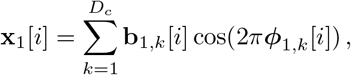

where 1 ≤ *i* ≤ *N*, and **b**_1,*k*_, ***ϕ***_1,*k*_ ∈ ℝ^*N*^ are uniformly sampled from *b*_1,*k*_(*t*) and *ϕ*_1,*k*_(*t*). Numerically, the estimation of **b**_1,*k*_, ***ϕ***_1,*k*_ and noise **n** from the PPG signal is achieved by the reconstruction formula for SST [5]. Then, compute `SQI`_M_every *d/f*_*s*_ s, where *d* ∈ ℕ is chosen by the user; that is,

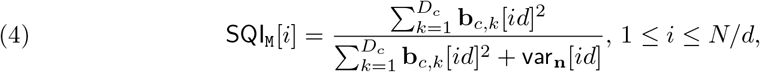

where

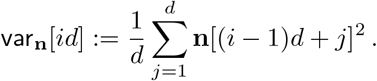

The Matlab implementation of `SQI`_M_ can be found in https://github.com/yanweiSu/PPG-SQIm.

We also compare `SQI`_M_ with existing SQIs, including the skewness (SQI_S_) [12, 8] computed for each 4 s PPG segment, the entropy (`SQI`_E_) [24, 8] for each 4 s PPG, and harmonic integrity index of order *n* ∈ ℕ, *H*_*n*_, motivated by studying the strength dynamics of various harmonics of ambulatory blood pressure signal (ABP) [30, 4]. Let **f**_1,*k*_ ∈ ℝ^*N*^ be the *k*-th harmonics component of the cardiac component, *H*_k_ of the *j*-th sample is defined as

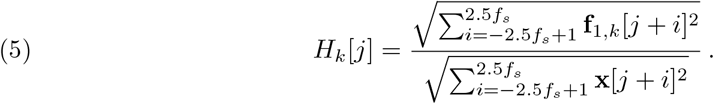

The perfusion index [8] is not considered since the databases we use have gone through a high pass filter.

### 3.1. Implementation details

Each PPG segment is 30 s in this paper. With *f*_*s*_ = 64 Hz, we used a 6th-order Butterworth bandpass filter with cutoff frequencies at 20 Hz and 0.5 Hz. Denote the pre-processed PPG segment as 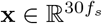. We used the second-order STFT-based synchrosqueezed transformation (SST) [18] with the window function 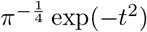, which leads to a *N* -by-*M* complex-valued matrix **S** ∈ ℂ^*N×M*^ as the discretized TFR, where *N* = 30*f*_*s*_, 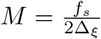, and Δ_*ξ*_ = 0.02 Hz. Then apply the multiple harmonics RD (MHRD) [26] on **S** with two ridges and parameters (*λ*_1_, *λ*_2_) = (1, 1) and (*µ*_1_, *µ*_2_) = (0, 0.07) to obtain IFs of the first two harmonics, followed by the single curve RD [26] with *λ* = 1 to obtain IFs of remaining higher harmonics, where we apply the masking technique; that is, at time *i/f*_*s*_, the band ranging from 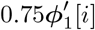 Hz to 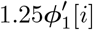 Hz is masked. Finally, set Δ = 0.2 to reconstruct *b*_1,*k*_(*t*) cos(2*πϕ*_1,*k*_ (*t*)), where *k* = 1, …, 5, denoted as **f**_1,*k*_ ∈ ℂ^*N*^, which leads to **b**_1,*k*_, ***ϕ***_1,*k*_ ∈ ℝ^*N*^, where **b**_1,*k*_[*i*] = |**f**_1,*k*_[*i*]| and ***ϕ***_1,*k*_ comes from phase unwrapping **f**_1,*k*_.

### 3.2. Train an interpretable signal quality assessment model

For each 30 s PPG segment, the label sequence is a {0, 1}-valued sequence, 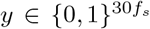, where 1 indicates “with artifact” (low quality) and 0 indicates “no-artifact” (high quality). To avoid the boundary effect, the first and last 5 seconds are discarded. This is not a serious problem since in practice the PPG signal is usually much longer than 30 second. To speed up, downsample *y* to 2Hz by the voting process over each 0.5s. The features defined by different SQIs are converted correspondingly by taking the median over each 0.5s. With SQIs and labels from all 30 second segments in the training dataset, we apply an interpretable learner, *Light Gradient Boosting Machine (LightGBM)* [11], to train a SQA model, with the learning rate of 0.1, the max number of leaves of each tree 7, the max number of bins for the feature values 255, and the cross-entropy as the loss function.

## 4. Materials and statistics

### 4.1. Dataset

We employed the publicly available dataset from [9] for validating the proposed SQA model. There are 7,306 segments with quality annotations in total. The labels are binary (1 for “artifact” or “low quality”, and 0 for “clean”, “no artifact” or “high quality”) to each sample point in the segment. These segments are derived from three public datasets: DaLiA [20], TROIKA [31], and WESAD [23]. Details of data preparation and labeling can be found in [9].

### 4.2. More details about the public databases

The employed publicly available datasets with experts’ labels [9] can be downloaded from https://github.com/chengstark/Segade/tree/main/data. There are 7,306 30-second PPG recordings in total, each accompanied by quality annotations. The labels assign binary values (1 for “artifact” or “low quality”, and 0 for “clean”, “no artifact” or “high quality”) to each sample point in the segment. These segments are derived from processing PPG signals from three public datasets: DaLiA [20], TROIKA [31], and WESAD [23], and the set that comes from DaLiA is split further into one training set, called the DaLiA-training (DTrain) set, and one testing set, called the DaLiA-testing (DTest) set.

All 30-second PPG segments are uniformly sampled at the sampling rate 64 Hz, and the signal values are normalized to the range [0, 1]. A second-order Butterworth filter with a low end cutoff of 0.9 Hz and a high end cutoff of 5 Hz was applied to the segments of both DaLiA and WESAD dataset^1^ by the authors in [9]. The TROIKA dataset was pre-processed by its original author in [31] with bandpass from 0.4 Hz to 5 Hz.^2^ To be consistent, we pre-process the databases used in [9] by applying a 6th-order Butterworth filter with a low end cutoff of 0.5 Hz and a high end cutoff of 20 Hz.

In the TROIKA dataset, both the PPG signal and triaxial acceleration signal were recorded from the wrist. Subjects performed treadmill running with changing speeds during data collection. For datasets labeled as `TYPE01`, running speeds changed as follows: rest (30 s) → 8 km/hr (1 minute) → 15 km/hr (1 minute) → 8 km/hr (1 minute) → 15 km/hr (1 minute) → rest (30 s). For datasets labeled as `TYPE02`, illustrated in Figure 1 in the main article, running speeds changed as follows: rest (30 s) → 6 km/hr (1 minute) → 12 km/hr (1 minute) → 6 km/hr (1 minute) → 12 km/hr (1 minute) → rest (30 s) [31]. In the DaLiA dataset, subjects performed 8 activity statuses plus one transient status: sitting, ascending and descending stairs, table soccer, cycling, car driving, lunch break, walking, and working, marked by IDs 1 to 8, respectively. The transient state, representing transitions between statuses, is marked by ID 0. Both the PPG signal used for analysis and the accelerometer signal plotted in Figure 1 in the main article were recorded from the wrist-worn device. Both TROIKA and DaLiA datasets provide ECG signals and the detected R peaks as ground truth for HR estimation. The WESAD dataset was recorded from both wrist- and chest-worn devices, from 15 subjects (age ranging from 21 to 55 years old, median 28 years old) during a lab study under different emotional states, including neutral, stress, and amusement. Subjects were allowed to move freely while performing tasks. The signals include ECG signals, tri-axis accelerometer signal, electrodermal activities record and PPG signals. The PPG signals in WESAD dataset are recorded from the wrist at the sampling rate 64 Hz [23].

The label generation procedure used in [9] is summarized here for readers’ convenience. Binary labels were created based on annotators’ observations of the three-axis acceleration signal, examining the correlation between ECG heartbeats and PPG heartbeats, and assessing the regularity of the PPG signals to identify artifacts. Two scenarios were considered for artifact annotations: (1) If the accelerometer shows motion and irregularities in the PPG signal align with the accelerometer data, the segment is marked as an artifact. (2) If the accelerometer shows no obvious motion, ECG displays a normal sinus rhythm, but irregularities are observed in the PPG signal, the segment is marked as an artifact. Each signal was annotated by at least one annotator. In the initial annotation trial phase, fifty 30-second segments were randomly selected and independently annotated by three annotators. Annotations from each pair of annotators were compared and analyzed, and the group of annotators collectively made decisions on correct annotations, improving agreement. The remaining data were annotated by a single annotator thereafter.

### 4.3. Learning process

We followed the procedure outlined in [9] to construct the training and testing sets. Specifically, 3436 segments from 12 subjects (ID 2 to ID 13) in the DaLiA dataset constitute the DaLiA-training (DTrain) set; 869 segments from the remaining subjects in the DaLiA dataset form the DaLiA-testing (Dtest) set. Additional testing sets include 2888 segments from the WESAD dataset and 113 segments from the TROIKA dataset. We allocate DaLiA-training for training and reserve DaLiA-testing, TROIKA, and WESAD for testing.

### 4.4. Statistical analysis

We first run a 10-fold cross-validation on the DTrain set following the 10-fold splitting in [9]. Then, train the SQA model on the entire DTrain set, and test on DTest, TROIKA and WESAD datasets separately. By viewing label 1 as the positive class, and 0 as the negative class, we report accuracy, sensitivity, precision, macro-F1 score, and the DICE score, which is defined as 2TP*/*(TP + FP + FN), where `TP, FP` and `FN` are true positive, false positive and false negative, respectively.

## 5. Result

In DTrain (DTest, TROIKA and WESAD respectively), the overall length of the PPG signal is 103,080 s (26,070 s, 3,390 s and 86,640 s respectively) and the overall length of artifact is 60,478.48 s (13,298.47 s, 1,784.58 s and 43,020.41 s respectively). Among all recordings, the ratio of labeled artifact in each recording is 0.59*±*0.34 (0.51*±*0.31, 0.53*±*0.34 and 0.50 *±* 0.39 respectively).

### 5.1. Basic statistics for signal quality indices

The mean and standard deviation of different SQIs are reported in Table 1. Overall, `SQI`_M_ and SQI_S_ are higher when the signal quality is high, which fits our expectation. *H*_1_ and *H*_6_ have opposite behavior, which can be explained by the fact that the higher order harmonic in PPG is weaker, and hence easily perturbed and “enhanced” by the high frequency component of artifacts.

**Table 1.** The mean and standard deviation of different SQIs.

| SQI | $y$ | DTrain | DTest | TROIKA | WESAD |
| --- | --- | --- | --- | --- | --- |
| $SQI_M$ | 1 | $0.85 \pm 0.10$ | $0.86 \pm 0.09$ | $0.85 \pm 0.11$ | $0.87 \pm 0.09$ |
| | 0 | $0.95 \pm 0.06$ | $0.95 \pm 0.06$ | $0.91 \pm 0.08$ | $0.98 \pm 0.03$ |
| $SQI_S$ | 1 | $0.10 \pm 0.60$ | $0.09 \pm 0.57$ | $0.07 \pm 0.54$ | $0.16 \pm 0.72$ |
| | 0 | $0.61 \pm 0.45$ | $0.62 \pm 0.47$ | $0.30 \pm 0.35$ | $0.77 \pm 0.44$ |
| $SQI_E$ | 1 | $1.46 \pm 0.46$ | $1.58 \pm 0.40$ | $1.68 \pm 0.33$ | $3.18 \pm 0.63$ |
| | 0 | $1.53 \pm 0.44$ | $1.53 \pm 0.42$ | $1.86 \pm 0.26$ | $3.20 \pm 0.42$ |
| $H_1$ | 1 | $0.54 \pm 0.14$ | $0.56 \pm 0.14$ | $0.52 \pm 0.15$ | $0.57 \pm 0.16$ |
| | 0 | $0.73 \pm 0.13$ | $0.74 \pm 0.13$ | $0.62 \pm 0.21$ | $0.78 \pm 0.10$ |
| $100H_6$ | 1 | $1.26 \pm 1.88$ | $1.02 \pm 1.39$ | $1.13 \pm 1.25$ | $1.30 \pm 1.70$ |
| | 0 | $0.57 \pm 0.97$ | $0.64 \pm 0.91$ | $0.65 \pm 0.87$ | $0.67 \pm 0.93$ |

### 5.2. Performance of each SQI

The Wilcoxon rank sum test on each testing dataset shows that all SQIs are significantly different (*p <* 10^−10^) on the artifact and the non-artifact groups. In DTrain (DTest, TROIKA and WESAD respectively), the Pearson correlation coefficients between `SQI`_M_ and *H*_1_, *H*_6_, SQI_S_ and SQI_E_ are 0.64, 0.07, 0.36 and −0.04 (0.59, 0.13, 0.40 and −0.09, 0.27, 0.24, 0.04 and 0.23, and 0.72, −0.04, 0.43 and −0.14 respectively) respectively. Except for TROIKA, the correlation coefficient between `SQI`_M_ and *H*_1_ is usually higher than 0.5. The area under the receiver operating characteristic curve (AUROC) and optimal threshold for the binary classification are reported in Table 2. Overall, except for TROIKA, the AUROC of `SQI`_M_ is the highest, and those of *H*_1_ and SQI_S_ are also high. Signals in TROIKA were recorded during running and were expected to be more challenging. Since `SQI`_M_ has the highest AUROC in general, we evaluate its ability as a single index to classify the signal quality. First, we learn the optimal threshold of `SQI`_M_ from the AUROC from DTrain using the experts’ labels. Then apply this threshold to the testing databases. Overall, the accuracy, macro-F1 and DICE are 0.78, 0.77 and 0.80 (0.64, 0.61 and 0.72, 0.85, 0.85 and 0.85, respectively) for DTest (TROIKA and WESAD, respectively).

**Table 2.** AUROC and the best thresholds of each feature for each testing datasets. The negative sign preceding an index emerges when the AUROC with the original index is below 0.5, prompting us to invert the sign of the index and report the resulting AUROC.

| **DTest** | $\text{SQI}_M$ | $\text{SQI}_S$ | $-\text{SQI}_E$ | $H_1$ | $-H_6$ |
| --- | --- | --- | --- | --- | --- |
| AUROC | 0.84 | 0.79 | 0.53 | 0.84 | 0.56 |
| Threshold | 0.96 | 0.46 | -1.39 | 0.69 | -0.01 |
| **TROIKA** | $\text{SQI}_M$ | $\text{SQI}_S$ | $\text{SQI}_E$ | $H_1$ | $-H_6$ |
| AUROC | 0.69 | 0.64 | 0.67 | 0.68 | 0.69 |
| Threshold | 0.94 | 0.15 | 1.92 | 0.68 | -0.003 |
| **WESAD** | $\text{SQI}_M$ | $\text{SQI}_S$ | $-\text{SQI}_E$ | $H_1$ | $-H_6$ |
| AUROC | 0.93 | 0.80 | 0.54 | 0.88 | 0.62 |
| Threshold | 0.96 | 0.45 | -3.58 | 0.71 | -0.01 |

**Table 3.** The confusion matrices and the performance metrics of the trained SQA model on different testing sets. NPV: negative predictive value; SEN: sensitivity; SPE: specificity; PRE: precision; mF1: macro-F1.

| **DTest** | 1 (Prediction) | 0 (Prediction) |
| --- | --- | --- |
| 1 (Label) | 15939 | 1984 |
| 0 (Label) | 4114 | 12723 |
| SEN = 0.89 | PRE = 0.80 | F1 (label 1) = 0.84 |
| SPE = 0.76 | NPV = 0.87 | F1 (label 0) = 0.81 |
| accuracy = 0.83; mF1 = 0.82 |  |  |
| **TROIKA** | 1 (Prediction) | 0 (Prediction) |
| 1 (Label) | 2200 | 198 |
| 0 (Label) | 901 | 1221 |
| SEN = 0.92 | PRE = 0.71 | F1 (label 1) = 0.80 |
| SPE = 0.58 | NPV = 0.86 | F1 (label 0) = 0.69 |
| accuracy = 0.76; mF1 = 0.75 |  |  |
| **WESAD** | 1 (Prediction) | 0 (Prediction) |
| 1 (Label) | 48902 | 8115 |
| 0 (Label) | 6783 | 51720 |
| SEN = 0.86 | PRE = 0.88 | F1 (label 1) = 0.87 |
| SPE = 0.88 | NPV = 0.86 | F1 (label 0) = 0.87 |
| accuracy = 0.87; mF1 = 0.87 |  |  |

### 5.3. Performance of the SQA model

The proposed SQA model achieves accuracy 0.86 *±* 0.01 and macro-F1 score 0.85 *±* 0.01 on DTrain under the 10-folds cross-validation scheme. When the trained model is tested on DTest (TROIKA and WESAD respectively), it achieves accuracy 0.83 (0.76 and 0.87 respectively), macro-F1 score 0.82 (0.75 and 0.87 respectively). See Table 6 for details. Note that DICE does not outperform the neural network based algorithm proposed in [9], which achieves 0.87, 0.81 and 0.91 in DTest, TROIKA and WESAD respectively, and we will come back to this in Discussion.

### 5.4. More analysis results

The histogram and receiver operating characteristic curve (ROC) of various SQIs over different databases are shown in Figures 2, 3 and 4.

**Figure 2.**
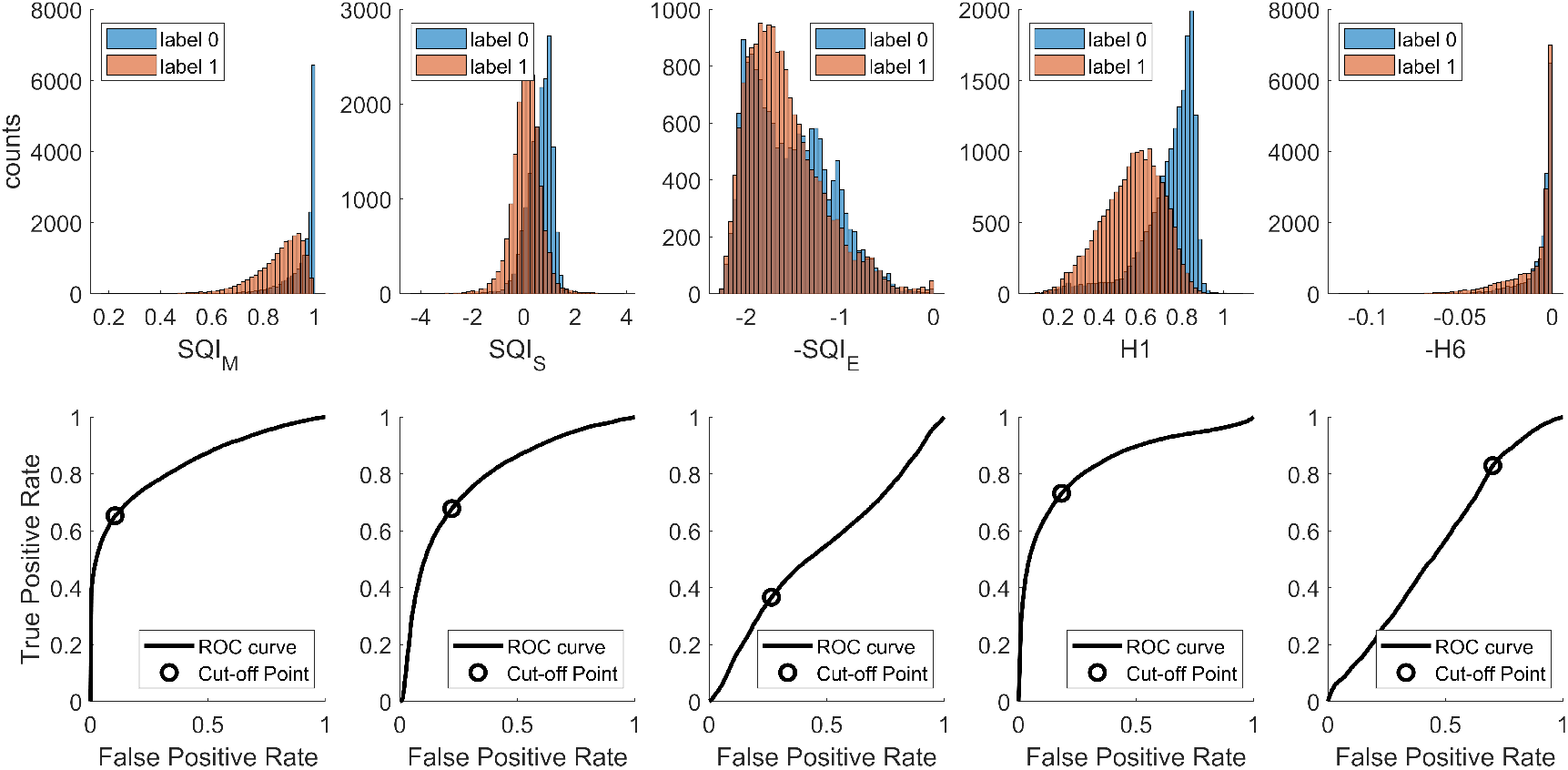
Histograms and AUROC curves of different SQIs on the DaLiA- testing dataset.

**Figure 3.**
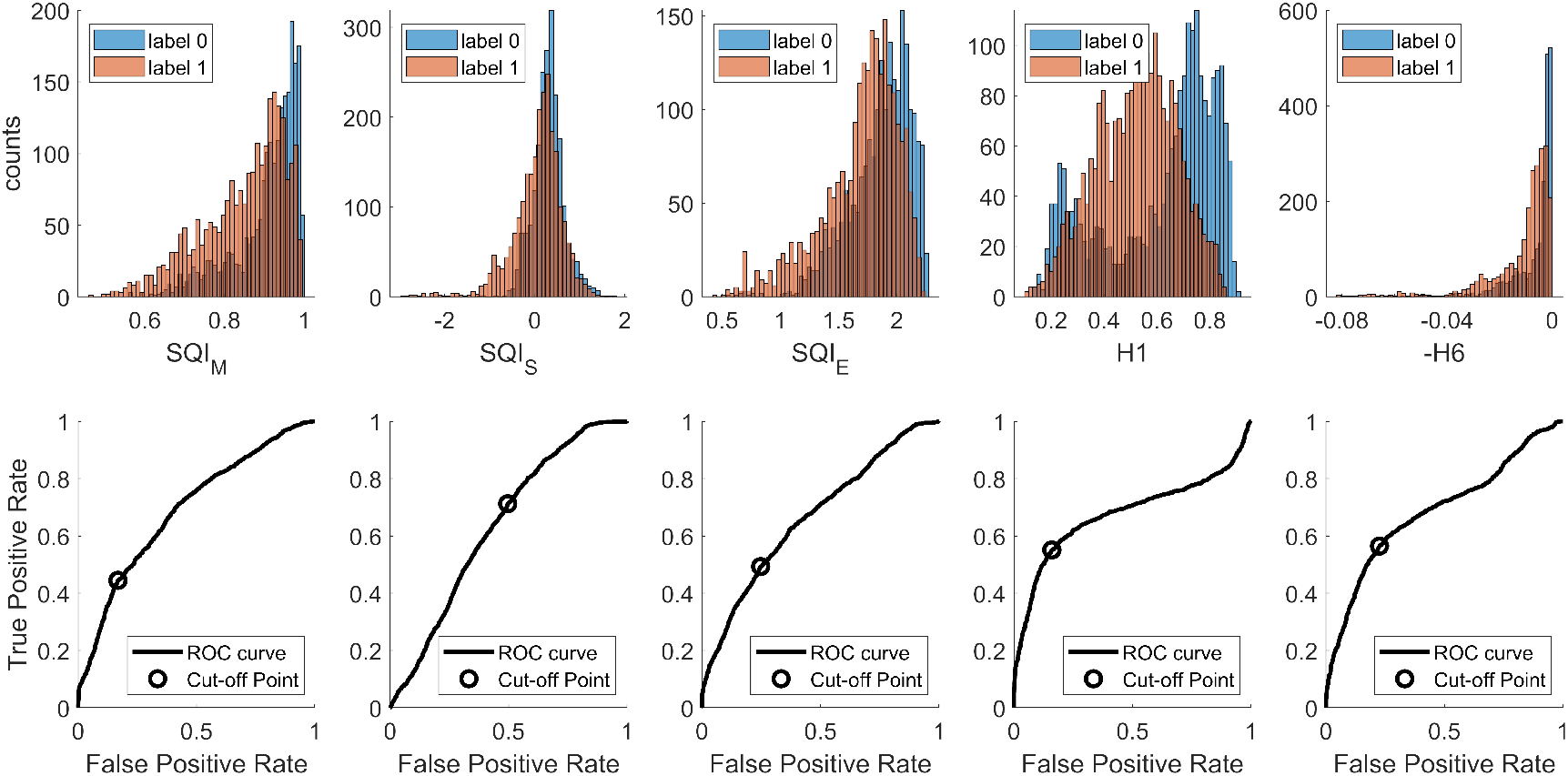
Histograms and AUROC curves of different SQIs on the TROIKA dataset.

**Figure 4.**
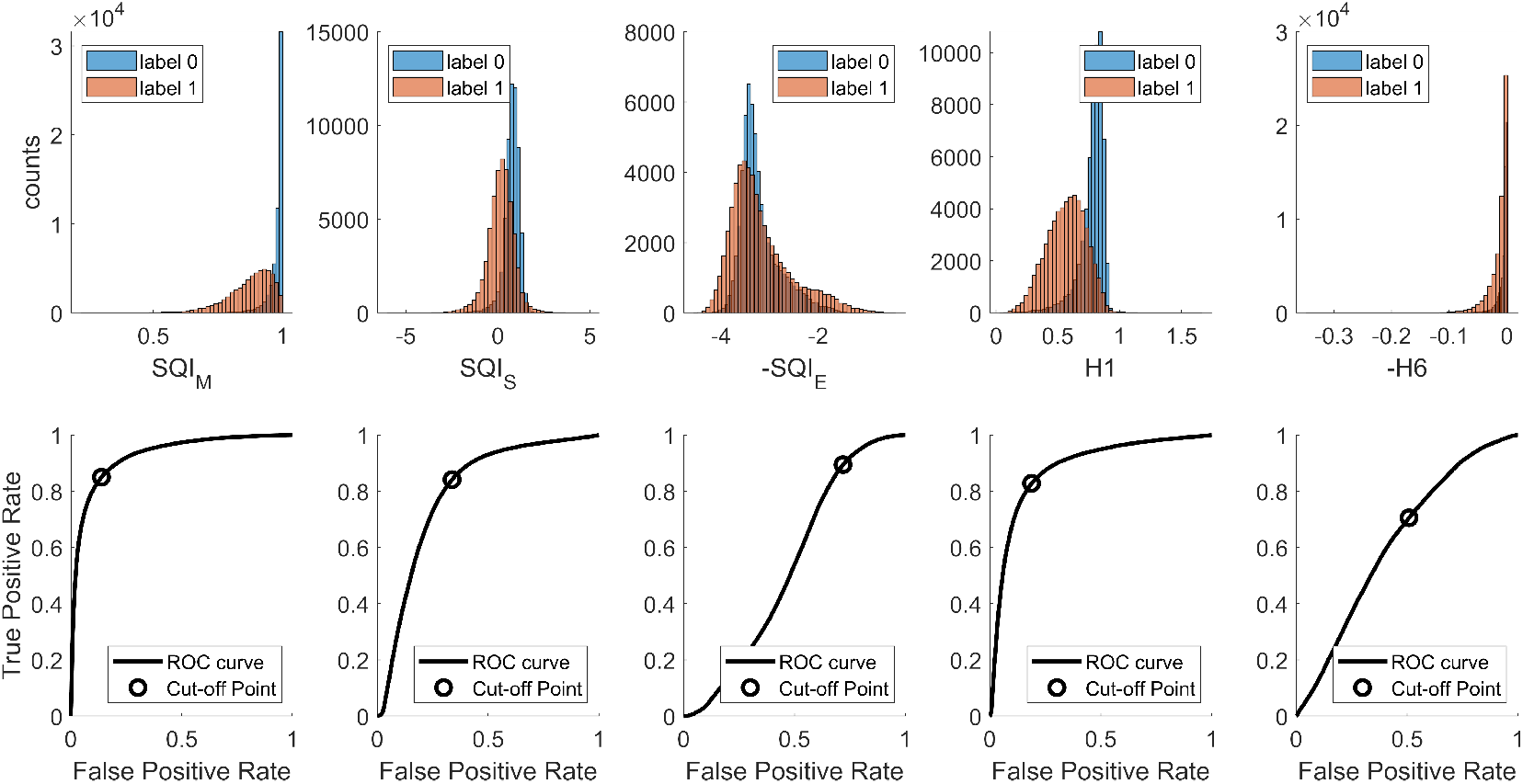
Histograms and AUROC curves of different SQIs on the WE- SAD dataset.

**Figure 5.**
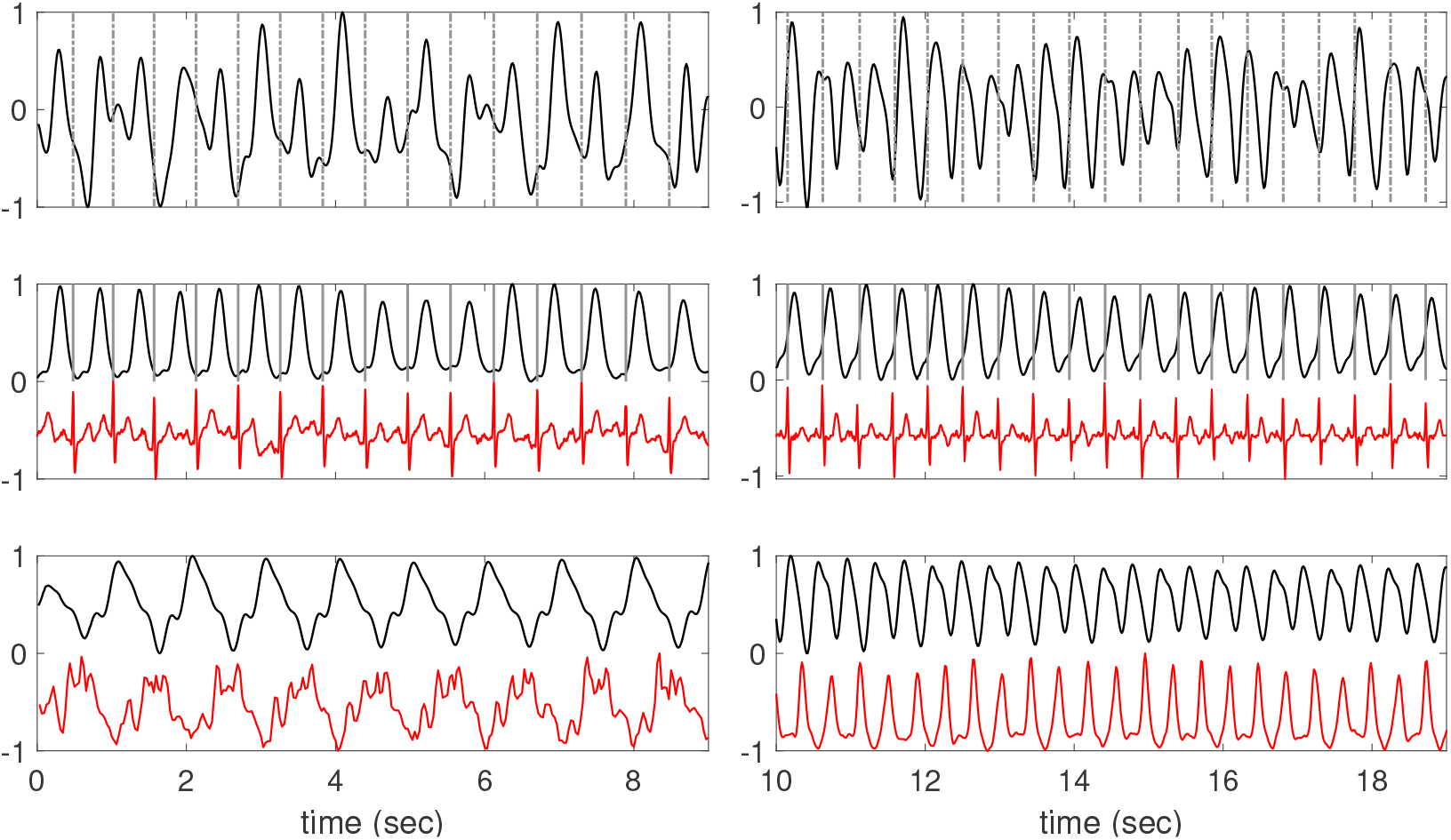
Two PPG signals recorded from two subjects’ wrists while they were running. Top row: the raw PPG signal that has been bandpass filtered with the 0.4 − 5Hz band. Middle row: the cardiac component decomposed from the raw PPG signal is shown as the black curve, and the simultaneously recorded ECG signal and the detected R-peaks are shown as the red curve and the grey lines, respectively. Bottom row: the motion rhythm decomposed from the raw PPG signal is shown as the black curve, and the magnitude of the simultaneously recorded accelerometer signal is shown as the red curve.

Among various SQIs, since `SQI`_M_ has the highest AUROC in general, we evaluate its ability as a single index to classify the signal quality in different databases. First, we learn the optimal threshold of `SQI`_M_ from the AUROC curve from DTrain using the experts’ labels. We then apply this threshold to DTest, TROIKA and WESAD. The result is shown in Table 4. Overall, accuracy and macro-F1 are 0.78 and 0.77 (0.64 and 0.61, 0.85 and 0.85, respectively) for DTest (TROIKA and WESAD, respectively).

**Table 4.**
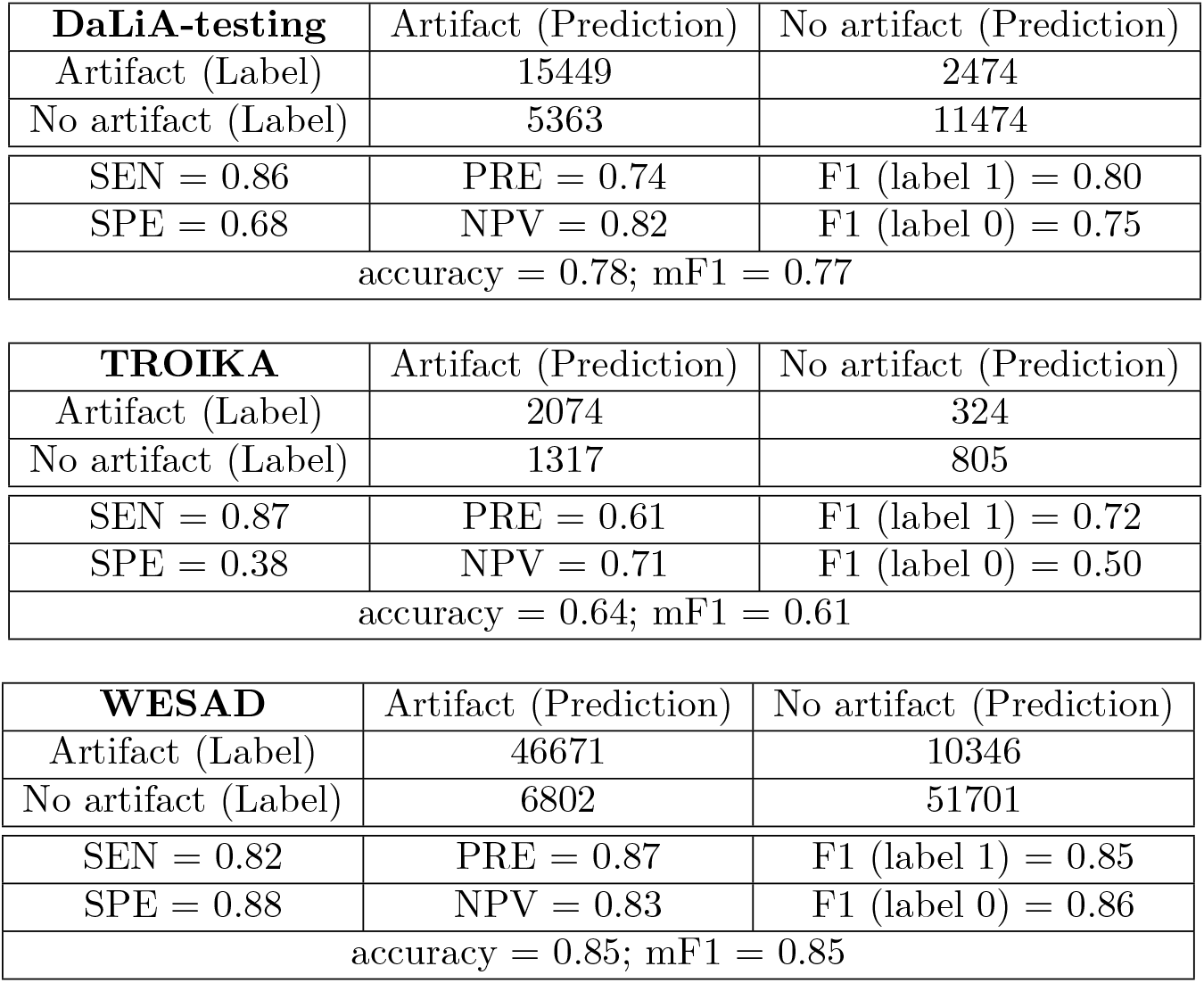
Performance evaluation of SQI_M_. We apply the threshold determined by DaLiA-training set on the other three testing datasets, and report the confusion matrices and standard metrics. NPV: negative predictive value; SEN: sensitivity; SPE: specificity; PRE: precision; mF1: macro- F1.

The proposed SQA model achieves accuracy 0.86 *±* 0.01 and macro-F1 score 0.85 *±* 0.01 on DTrain under the 10-folds cross-validation scheme, which is shown in Table 5. We follow the 10-fold splitting proposed in [9].

**Table 5.**
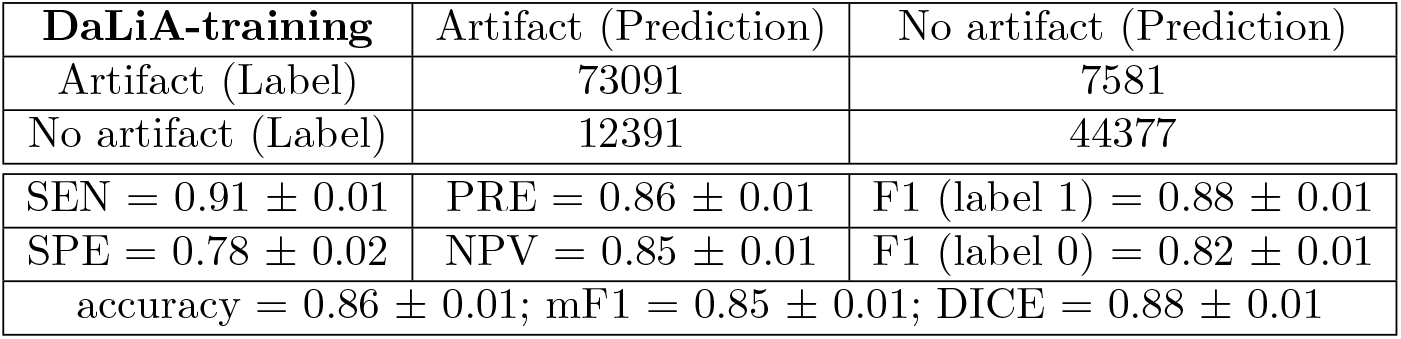
The 10-folds cross-validation of the proposed SQA model on DaLiA-training set. The total sum of 10 confusion matrices is shown above. NPV: negative predictive value; SEN: sensitivity; SPE: specificity; PRE: precision; mF1: macro-F1.

**Table 6.**
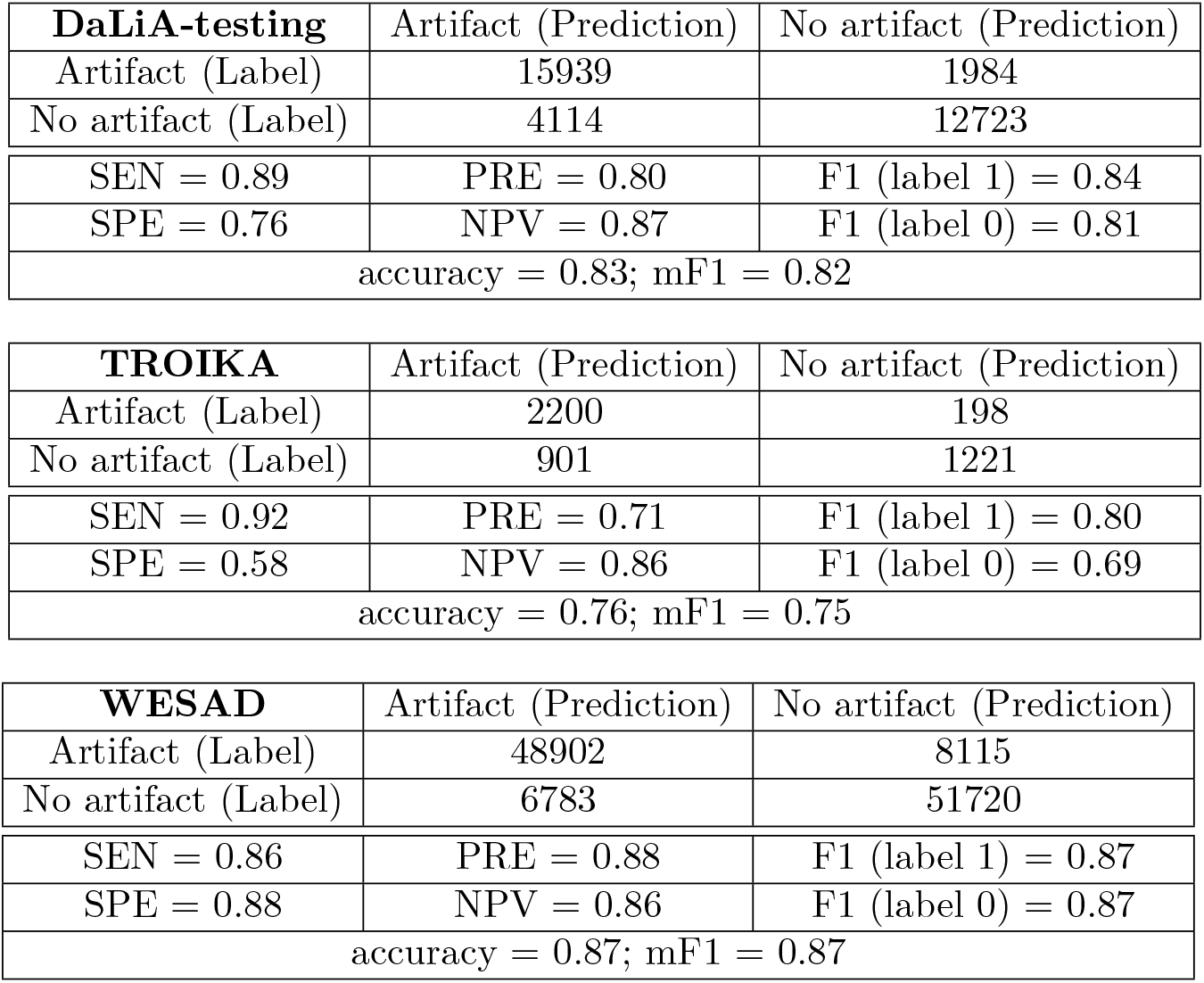
Sum of the confusion matrices and the performance metrics of testing the SQA models on each testing sets. The SQA model is trained from the DaLiA-training database. NPV: negative predictive value; SEN: sensitivity; SPE: specificity; PRE: precision; mF1: macro-F1.

When the trained model is tested on DTest (TROIKA and WESAD respectively), it achieves accuracy 0.83 (0.76 and 0.87 respectively) and macro-F1 score 0.82 (0.75 and 0.87 respectively). See Table 6 for details.

Finally, we compare the performance of the proposed signal quality indices with the **Segade** model proposed in [9]. The DICE scores of the proposed SQA model, SQI_M_, and **Segade** are reported in Table 7, where the DICE score is defined as `2TP`*/*(TP + FP + FN), where TP means true positive, FP means false positive and FN means false negative [9].

**Table 7.**
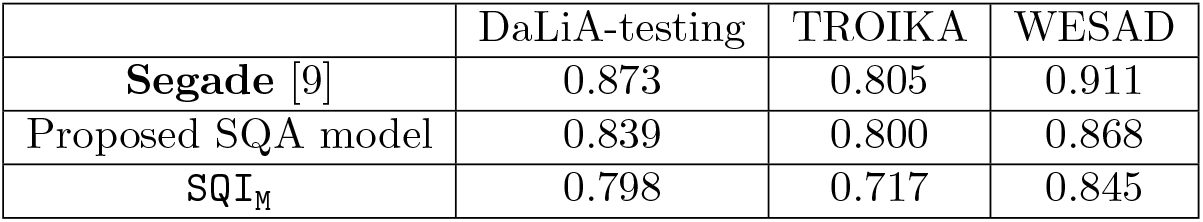
The DICE score of testing the proposed SQA model on each dataset. The **Segade** result in the first row is from Table 1 in [9].

## 6. Discussion and conclusion

We proposed a model-based SQI, denoted as SQI_M_, and a learning-based SQA model that incorporates various SQIs including SQI_M_. The proposed SQA performs well, but does not outperform the existing CNN-based approach model.

The first topic to discuss, which probably is the spotlight of readers interested in the “predictive model”, is the performance of our SQA model. In SQI_M_, the strong motion rhythm resistant to the bandpass filter is treated as “noise”, resulting in a small SQI_M_. This, coupled with concerns about labels derived from raw PPG signals raised in [27] elucidates the slightly lower performance of our SQA model compared to results reported in [9], which is a convolutional neural network model derived from the U-Net model architecture [21] tailored for 1D signal processing. See the left subplot of Figure 5 for a PPG segment that is labeled “low quality”, where the PPG is composed of a cardiac component and a motion rhythm since the subject was running at the speed of 6km/hour. This segment was considered of low quality, probably due to its irregular pattern, but its decomposed cardiac component is reasonably well. In the right subplot of Figure 5, the PPG segment is labeled “high quality” probably since its presence “seems” regular and close to cardiac oscillation. However, these cycles do not aligned with the cardiac cycles confirmed by the simultaneously recorded electrocardiogram (ECG). We thus could reasonably view the labeled signal quality as *uncertain*. This uncertainty complicates the comparison of model performances. Although the DICE evaluation of the proposed SQA model indicates a lower performance than [9], the SQA model holds a distinct advantage in interpretability inherited from the PPG model. This raises the question of whether quantifying signal quality of cardiac component post-decomposition is more effective when irrelevant components exist. As no labeled database follows this approach, we leave this intriguing question for future research.

The advantage that `SQI`_M_ is defined with mathematical meanings allows generalization for quantifying other information in PPG; for example, respiratory information, motion rhythm, or other factors in PPG signals. This is related to the change point detection for oscillatory signals in statistics, which unfortunately has received limited attention, except for recent efforts [29]. Note that respiratory information like RIIV may be absent, motion rhythms might be absent or irregular, and arrhythmia might appear, making quality assessment vague. We may extend the change point detection algorithm [29] to the PPG signal, considering time-varying frequency, amplitude, and WSF. This problem is prevalent in other scientific fields, and exploring joint oscillatory component change point detection and signal decomposition is a future research direction.

In conclusion, our proposed ANHM model, in conjunction with advanced signal decomposition tools, holds promise for establishing such a system by incorporating the signal decomposition step. With labels provided by this system, we can advance towards establishing a more dependable SQA model, particularly for scientific research.

## Data Availability

All data produced are available online and cited in the paper.

## Acknowledgement

The authors thanks author in [9] for providing details regarding the labeled databases they shared.

## Footnotes

1 https://github.com/chengstark/Segade/blob/main/db2npy.py, line 23 to line 30.

2 See [9] and the description in [31]

